# Optimal epidemic control under uncertainty: trade-offs between information collection and other actions

**DOI:** 10.1101/2022.03.28.22273039

**Authors:** Julien Flaig, Nicolas Houy

**Author notes:** Corresponding author. Epidemiology and Modelling of Infectious Diseases (EPIMOD), F-69002 Lyon, France. University of Lyon, Lyon, F-69007, France; CNRS, GATE Lyon Saint-Etienne, F-69130, France.

## Abstract

**Background:** Recent epidemics and measures taken to control them – through vaccination or other actions – have highlighted the role and importance of uncertainty in public health. There is generally a trade-off between information collection and other uses of resources. Whether this trade-off is solved explicitly or implicitly, the concept of value of information is central in order to inform policy makers in an uncertain environment.

**Method:** We use a deterministic SIR disease emergence and transmission model with vaccination that can be administered as one or two doses. The disease parameters and vaccine characteristics are uncertain. We study the trade-offs between information acquisition and two other measures: bringing vaccination forward, and acquiring more vaccine doses. To do this, we quantify the expected value of perfect information (EVPI) under different constraints faced by public health authorities, *i*.*e*. the time of the vaccination campaign implementation and the number of vaccine doses available.

**Results:** We discuss the appropriateness of different responses under uncertainty. We show that in some cases, vaccinating later or with less vaccine doses but more information about the epidemic and the efficacy of control strategies may bring better results than vaccinating earlier or with more doses and less information respectively

**Conclusion:** In the present methodological paper, we show in an abstract setting how clearly defining and treating the trade-off between information acquisition and the relaxation of constraints can improve public health decision making.

**Highlights:**

- Uncertainties can seriously hinder epidemic control, but resolving them is costly. Thus there are trade-offs between information collection and alternative uses of resources.
- We use a generic SIR model with vaccination and a value of information framework to explore these trade-offs.
- We show in which cases vaccinating later with more information about the epidemic and the efficacy of control measures may be better – or not – than vaccinating earlier with less information.
- We show in which cases vaccinating with less vaccine doses and more information about the epidemic and the efficacy of control measures may be better – or not – than vaccinating with more doses and less information.

## 1 Introduction

Infectious disease control is steeped in uncertainty. Uncertainty may concern the natural history of the pathogen, the successful development of prophylactic or curative treatments, the ability to deliver control interventions, or the population characteristics, including contact patterns and individual behaviors [1]. Since the spread of an infectious disease is a dynamic and nonlinear phenomenon, uncertainties can seriously hinder control efforts. Control measures taken at a certain time given available information may prove sub-optimal as more information is gathered about the epidemic, but these measures are usually irreversible and their cost is sunk.

Yet resolving uncertainties has a cost, so there is a *trade-off* between i) using resources (money and time) to gain information about the uncertain parameters of an epidemic, and ii) allocating the same resources to alternative uses meant to control that epidemic. Alternative uses of resources include the production of more medical supplies and the build-up of intervention capacity in general. They also include accelerating or bringing interventions forward in time, it being understood that since epidemics spread over time, acting swiftly will avoid further infection cases. How, then, to balance the benefit of information collection against the benefits of these alternative resource uses – that is, better informed decisions against faster and stronger interventions based on less information? The objective of this article is to show how this can be done through value of information analyses, and to provide a general understanding of these trade-offs.

Value of information analyses allow to quantify the benefit of reducing uncertainties. They are widely used in the fields of public health and health economics [2, 3]. One popular value of information metric is the expected value of perfect information (EVPI). The outcome of a decision (in our case: an epidemic control strategy) typically depends on both the decision itself, and the parameters that may be uncertain. EVPI is the difference between i) the expected outcome obtained by making decisions knowing the true value of all parameters, and ii) the expected outcome obtained by making decisions without knowing the true values of uncertain parameters. Put differently, EVPI is the expected additional value, given prior beliefs about the possible values of uncertain parameters, of making decisions knowing the true value of these parameters.

Of course, the two terms of EVPI (the outcomes with and without additional information) depend on *feasible* epidemic control strategies, that is on the constraints imposed on decision making. Whether decisions are made with or without knowing the true parameter values, the achievable best policy depends obviously on how fast and how broadly or strongly control measures *can* be implemented. Feasible strategies, in turn, depend on resource allocation: if, say, more medical supplies are produced, decision making will be less constrained and will result in a better (or equivalent) outcome. Hence, value of information analyses allow to compare the outcomes obtained with or without additional information and with or without accelerating or strengthening control measures, that is to balance the benefit of information collection against that of alternative uses of resources.

It should be kept in mind, however, that while EVPI is a popular metric allowing to readily and consistently quantify the value of information, this metric only provides an upper bound on the incremental benefit of collecting more information about uncertain parameters – this is because it assumes that perfect knowledge of parameters is achievable, which may of course not be the case. Other value of information metrics are useful in practice, in particular the expected value of sample information (EVSI), that is the expected value of observing an additional sample of uncertain parameters. We leave EVSI outside the scope of this article because it requires to actually define samples for each parameter (their nature, size, method of collection), which is likely to be very case specific and at any rate irrelevant to our point. All effects shown in this article will remain in an EVSI based analysis, although with reduced magnitude. Yet EVPI does not address one relevant issue, which is to determine what parameters are driving uncertainty i.e. those for which resolution of uncertainty would be the most beneficial. Doing so is useful to set information collection priorities. Hence, as a complement to EVPI, we also compute the expected value of partially perfect information (EVPPI) for uncertain parameters. EVPPI measures the additional benefit of knowing the true value of a subset of uncertain parameters. For instance, the additional benefit of knowing the value of one of the parameters, the others remaining uncertain.

In the present conceptual and methodological article, we use a generic deterministic SIR transmission model, with uncertain parameters, of an infectious disease emerging in a population. The control intervention is vaccination, and the decision maker can choose how many single and double dose vaccines to administer. The value of a vaccination policy is the number of averted symptomatic infected individual-days over the *entire* course of the epidemic compared to the no intervention scenario. We want to illustrate, first, how vaccinating earlier with less information compares to vaccinating later with more information and, second, how vaccinating with more vaccine doses but less information compares to vaccinating with less doses and more information.

Uncertainty management and information collection are ubiquitous issues in public health. In the present study, however, information collection is the result of active decision making, and uncertainty management is therefore a strategic issue. This needs to be contrasted with the way uncertainty is most often managed in infectious disease control studies.

It should first be noted that policy-oriented studies in the field of infectious disease control rarely consider the value of acquiring more information (see [4] for an example of study which does), and considering information collection as a strategic choice among others is even less common. Most studies focus on available information only and disregard the possibility of resolving uncertainties. A typical approach consists in using available data to fit a disease transmission model. This task can be resource intensive and is the main contribution of many studies. It results in estimates of the uncertain parameters (point estimates or distribution estimates depending on the method used). In a second step, these estimates are used to determine the best control policy either by comparing all relevant options exhaustively or via optimization methods. Finally, uncertainties are managed in a third step: the robustness of the chosen policy is tested by performing sensitivity analyses. The objective of these sensitivity analyses is not so much to quantify uncertainties (e.g. by using value of information metrics to estimate the benefit of reducing them) as to justify the choice of a policy based on available data and the corresponding model fit. Available information is assumed to be fixed and is used both to pick a policy, and to derive scenarios in which to test the performance of that policy.^1^ Even considering the objective of justifying a policy given available information, sensitivity analyses can easily be incomplete (limited sampling of the parameter space) or use misleading metrics (e.g. the probability that a given policy will be the best instead of the expected value of that policy). We refer to Houy and Flaig [5] for a discussion of these issues and examples from the field of hospital epidemiology.

Other studies assume that more information is acquired over time and propose algorithms to estimate parameters (see [6] and the articles reviewed therein) or to adapt interventions (see [7, 8], for instance) in real-time. Yet in these studies just as in studies assuming fixed information, information acquisition is seen as a *passive* process, and is not stemming from *active* strategic decisions. Namely, there is no trade-off between data collection and other uses of resources. Data is simply assumed to become available “for free” over time, and the point of these articles is to show how to use it. In this respect, whether information is assumed to be available at one point in time or to become available over time makes little conceptual difference.

To some extent, testing or screening optimization could be seen as strategic (or active) acquisition of information, however not in the sense of solving the trade-offs discussed in this article, as testing optimization does not entail assessing the value of information (at least not explicitly). In the context of infectious disease control, the objective of testing optimization is usually to control the spread of a disease by finding and isolating infectious individuals [9–13] or, less frequently, by finding recovered and immune individuals to end their isolation [14, 15]. Framing the problem in terms of value of information, the uncertain parameter would be the health statuses (susceptible, infectious, or recovered) of individuals. Yet the decision made once this information is available is typically predefined and fixed: it often simply consists in, say, isolating tested individuals found to be infectious, or in following a more sophisticated threshold-based rule by isolating a tested individual based also on other individuals’ test results. In both cases, the problem is to find *how* to test (who and when) given that such or such predefined decision (isolate or not) rule is applied depending on test results. By contrast, a value of information analysis would rather address the issue of *whether* to test or not given that test information could be used to make better decisions.

Our work is more closely related to several previously published studies. Cipriano and Weber [16] solved the problem of deciding when to stop screening successive cohorts of 50-year-olds for an infectious disease, hepatitis C, and when to optimally collect additional data to inform this decision. Their objective, however, was not to control the spread of the disease but to find cases. They did not model hepatitis C transmission and assumed that the prevalence decreased linearly over time. Closer to our work, some authors proposed multi-stage control strategies allowing to actively include information collection in decision making rather than passively adapt to new information. In the first example developed in their study, Shea et al. [17] showed how anticipating that uncertainty will be resolved at a later stage can influence decisions in earlier stages. The trade-off between acting early or collecting information and acting later is solved implicitly, but it is not the main focus of the article – the trade-off is solved for a single late (informed) intervention date. Similarly, Atkins et al. [18] considered a scenario in which the efficacy of a vaccine is uncertain but can be learned through vaccination campaigns. In this study as well, the focus is on the comparison of active information collection versus passive adaptation, and trade-offs are solved implicitly. The trade-off between trying to control an epidemic early with less information and later with more information is treated explicitly and in more details by Thompson et al. [19]. However they did not used an EVPI framework but proposed an heuristic algorithm allowing to anticipate future learning. We want to bring the discussion to a more general and elemental level. The relationship between logistical constraints (or equivalently resource or budget constraints) and the value of information was investigated by Shea et al. [17] (second example of the paper) and by Li et al. [20], however not in terms of the trade-off between relaxing these constraints and collecting information about the epidemic. This trade-off is illustrated by Woods et al. [21] in the case of HIV, but not for the full range of resource constraints. In this article, we address both implementation time and resource constraints and focus on trade-offs more in-depth.

The remainder of the paper is structured as follows. In Section 2 we introduce the SIR disease transmission model (Section 2.1), model uncertainties (Section 2.2), control interventions (Section 2.3), and the value of information metrics used in the article (Section 2.4). The simulation results are shown and discussed in Section 3. Sections 3.1 and 3.2 give preliminary results. The core results of the paper are presented in Section 3.3. Section 4 concludes.

## 2 Materials and methods

### 2.1 Disease transmission model

We consider the emergence of an infectious disease in a closed population of *N* = 10,000 individuals. The spread of the disease is described by a SIR model with symptomatic and asymptomatic infections, and vaccination. Individuals may receive 0, 1, or 2 doses of vaccine. Vaccination is assumed to reduce infectiousness, susceptibility, and the probability of symptomatic infection. For concision and illustrative purposes, we use a deterministic model (with continuous population) and leave aside stochastic effects, although they can be critical in the context of infectious disease emergence (see [19]). At time 0, no individual is vaccinated and one individuals out of *N* is assumed to be infected. Simulations are run until the epidemic is extinct. Since population is continuous in our framework, we need to define a threshold number of infectious individuals below which the epidemic is assumed to be extinct. We set this threshold to *N ×* 10^−8^ = 10^−4^ infectious individuals. In our scenario, extinction is reached after some time for any model parameter values because we assumed a SIR transmission model in a closed population.

The disease transmission model is illustrated in Figure 1 and the parameters are defined in Table 1. The model equations are given in Appendix A and the model parameters in Table 1. The unit of time is one day throughout the article.

**Figure 1:**
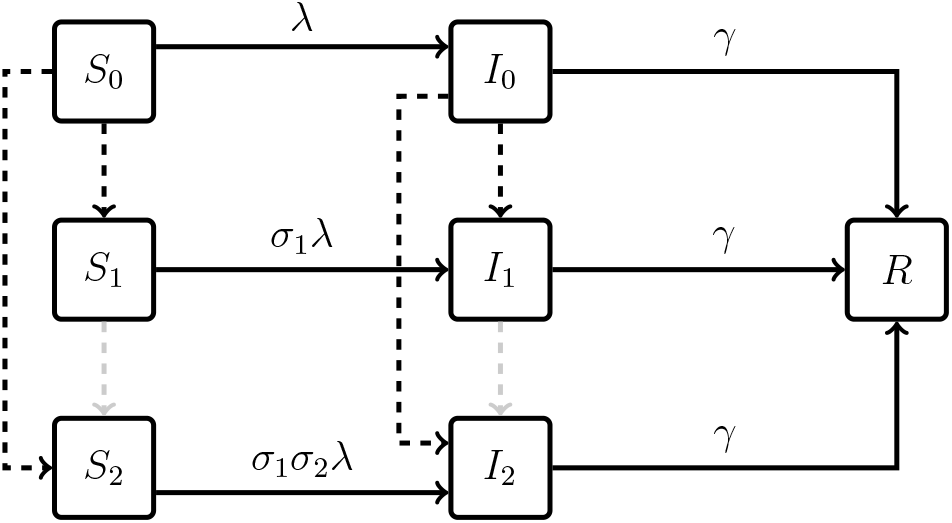
Sketch of the transmission model. Dashed: possible vaccination. Grey dashed: vaccination not considered in the present article (see vaccination strategies in section 2.3). *λ* is the force of infection for unvaccinated individuals.

**Table 1:** Transmission model parameters.

| Parameter | Description |
| --- | --- |
| $R_0$ | Reproduction number of unvaccinated individuals. |
| $\gamma$ | Rate of recovery. |
| $\rho_1$ | Relative infectiousness of individuals having received 1 dose of vaccine, compared to 0 dose. |
| $\rho_2$ | Relative infectiousness of individuals having received 2 doses of vaccine, compared to 1 dose. |
| $\sigma_1$ | Relative susceptibility of individuals having received 1 dose of vaccine, compared to 0 dose. |
| $\sigma_2$ | Relative susceptibility of individuals having received 2 doses of vaccine, compared to 1 dose. |
| $p_1$ | Probability of symptomatic infection of individuals having received 1 dose of vaccine. |
| $p_2$ | Reduction of the probability of symptomatic infection of individuals having received 2 doses of vaccine, compared to 1 dose. |

### 2.2 Uncertainties

We assume the model structure to be known and focus on parameter uncertainties. We also assume prior distributions to be available for all uncertain parameters. Prior distributions represent i) the *level of information* (i.e. of certainty) about parameter values, and ii) *expectations* about these values at the time of decision. We show results for a set of prior distributions which assumes that some information is available about the parameters (there is some variance, but some values are more likely than others), and that double dose vaccination is expected to provide markedly more protection than single dose vaccination. The details of baseline prior distributions are given in Appendix B and Figure 2 shows 10,000 draws from these distributions. We assume the rate of recovery *γ* to be known and equal to 0.1 day^-1^, so that all uncertainty about the spread of the disease without intervention is reflected by uncertainty about a single parameter, *R*_0_, which in our case is equivalent to an uncertain contact rate *β* = *γR*_0_. All other uncertain parameters concern the efficacy of one dose and double dose vaccination.

**Figure 2:**
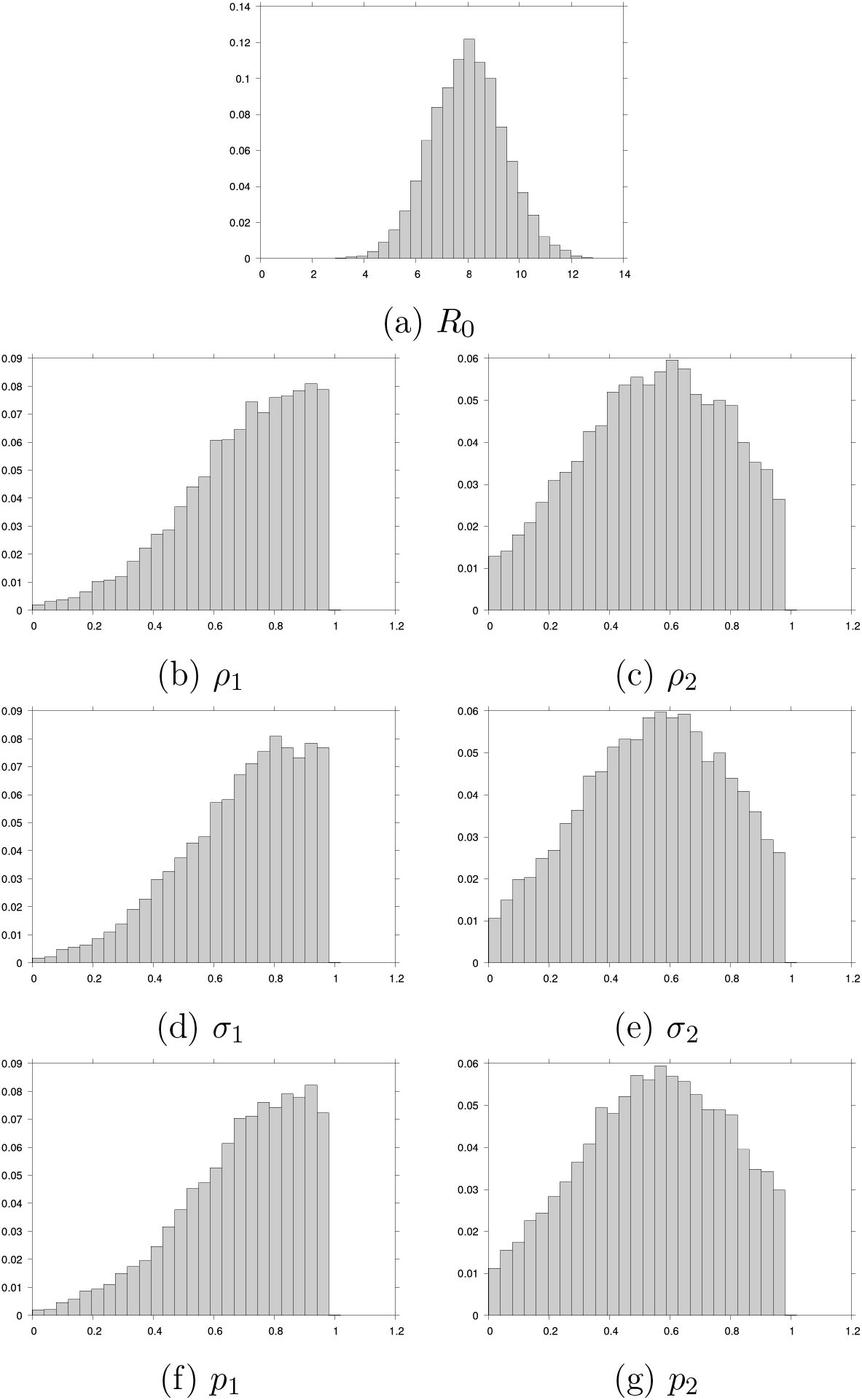
10,000 draws from the baseline prior distributions.

### 2.3 Optimization problem and vaccination strategies

Our objective is to vaccinate the population so as to minimize the total number of symptomatic infected individual-days over the entire course of the epidemic, that is from time 0 until the epidemic is extinct. We assume no time discounting.

In our scenario, a vaccination strategy consists in administering *n*_1_ vaccine doses as single doses, and *n*_2_ vaccine doses as double doses (i.e. to *n*_2_*/*2 individuals) to previously unvaccinated individuals at a given time *t*. The set of alternative vaccination strategies is constrained by i) the time *t*_min_ from which vaccine doses can be administered, and ii) the number of doses *n*_max_ available.

Clearly, *t*_min_ is the vaccination date since, other things being equal, vaccinating earlier is better than vaccinating later. In the following, we will alternatively refer to *t*_min_ as the vaccination date, and as a constraint on the vaccination date that may or may not be relaxed.

For concision, we assume that all *n*_max_ doses are available at once and administered at a single date by bundles of 2,000 doses (*n*_1_ and *n*_2_ are multiples of 2,000). We can expect that in real life, vaccine doses will be produced and administered at a certain rate, resulting in a gradual increase of coverage. Also, the assumption that the number of individuals actually getting vaccinated can be controlled loosely corresponds to a mandatory vaccination scenario. Finally, notice that we assume that individuals are vaccinated irrespective of their health status. In practice, vaccination campaigns could be made more efficient by using information on who is symptomatically infected and who recovered from a symptomatic infection and targeting individuals accordingly. In the following, we will denote the set of feasible vaccination strategies as *A*(*t*_min_, *n*_max_). A formal definition of *A*(*t*_min_, *n*_max_) is given in Appendix C.

Optimization involves deciding whether to vaccinate more individuals with a single dose, or less individuals with a double dose. Which option is better will depend on the values of uncertain parameters (the relative efficacy of single and double dose vaccination in particular) and on the set of feasible policies (*n*_max_ in particular).

### 2.4 Value of information

EVPI is computed as follow. Put generally, a decision-maker chooses a strategy *a* in a set *A* of alternatives to maximize a value function *V* (e.g. the number of averted cases or averted costs over the course of the epidemic) that also depends on uncertain parameters denoted *ξ*. Under uncertainty about the true value of *ξ*, an alternative *a*_0_ is usually picked that maximizes the expected value over possible realizations of 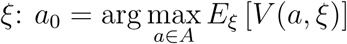. If there is no uncertainty and the true value *ξ*^*^ of *ξ* is known, an alternative *a*^*^ can be picked that simply maximizes the value *V* given 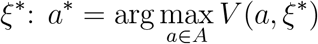. The difference 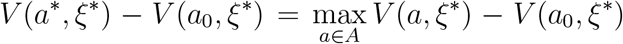 is the value loss due to making a decision under uncertainty. EVPI is the expected value loss over possible realizations of 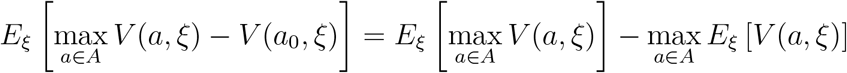. In other words, EVPI quantifies uncertainty as the expected benefit of resolving it, and hence gives an upper bound on the cost one should be willing to pay to reduce it.

A simple tweak allows to use an EVPI framework to balance information acquisition against other resource uses. The two terms of EVPI can be computed separately:

1. the *ex ante* maximum expected value 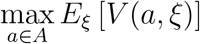, that is the expected value of controlling the epidemic without collecting more information (i.e. *before* the true value of *ξ* is known), and

2. the expected *ex post* maximum value 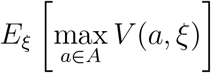, that is the expected value of controlling the epidemic with perfect knowledge of *ξ* (i.e. *after* the true value of *ξ* is known).

Using resources for other uses than collecting information may, for instance, broaden the set *A* of feasible policies (in our scenario, by decreasing *t*_min_ or increasing *n*_max_). Thus, computing the two terms of the EVPI for different sets *A* of feasible policies allows to explore the trade-off between collecting information and alternative uses of resources.

For each of the 10,000 draws of parameter values *ξ* (Figure 2), we compute the reduction in symptomatic individual-days *V* (*a, ξ*) allowed by each alternative policy in *A*(*t*_min_, *n*_max_). Then, we estimate the maximum ex ante expected value and the expected ex post maximum value of epidemic control, that is the value of resolving uncertainties given the set *A*(*t*_min_, *n*_max_) of feasible policies. By considering different values for *t*_min_ and *n*_max_, we show the trade-off between relaxing these constraints and resolving parameter uncertainties.

Like EVPI, EVPPI assumes that under uncertainty, a policy *a*_0_ is picked that maximizes the expected value over possible values of uncertain parameters. It is then assumed that uncertainties can be resolved for subsets of uncertain parameters, typically a single parameter. Let *θ* denote this subset of parameters. If the true value *θ*^*^ of *θ* is known, then policy 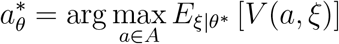 is picked. The value loss due to choosing a policy without knowing the true value of *θ* is 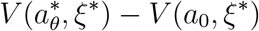. The EVPPI is the expected loss over possible values of 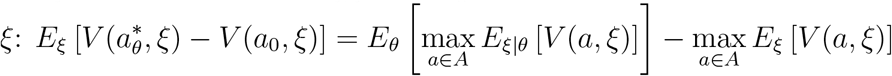.

EVPPI estimation is resource intensive. We estimate the expected value 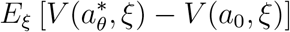 over 3,500 “true” realizations *ξ*^*^ of *ξ*. Policy *a*_0_ is picked based on 1,000 other realizations of *ξ*. Policy 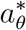 is picked based on the same 1,000 draws, but using the true value *θ*^*^ of *θ* (a subset of *ξ*^*^). Both EVPI and EVPPI are positive by definition. Notice that due to stochastic effects, EVPPI estimates can be negative or their confidence intervals include negative values, while this is not the case of EVPI estimates.

## 3 Results

### 3.1 Decision under uncertainty

Without vaccination, the expected number of symptomatic infected individual-days over possible parameter values is 99,926 with 95 % confidence interval (CI) 99,922.6 – 99,929.4. In our scenario, given a number of available vaccine doses *n*_max_ and a time from which vaccination is possible (i.e. a vaccination date in our case) *t*_min_, deciding on a vaccination policy consists in deciding on the number of doses to be administered as single and double doses. Double doses vaccination is expected to be more efficient, but in some cases vaccinating more people with a single dose might be optimal from a public health perspective.

Figure 3 shows the (ex ante) expected reduction in symptomatic infected individual-days as a function of the number of vaccine doses distributed as single doses, the remaining doses being distributed as double doses, for vaccination at time *t*_min_ = 0 and *n*_max_ = 10,000 available doses. Under uncertainty, in this case, the policy *a*_0_ maximizing expected value consists in administering all available 10,000 doses as single doses. This results in an expected 36,274.5 (95 %CI: 35,799.6 – 36,749.4) averted symptomatic infected individual-days on average, which is the ex ante maximum expected value. The worst policy consists in administering 4,000 single doses and the remaining 6,000 doses as double doses to 3,000 individuals, and the performance of administering 5,000 double doses (0 single doses) lies in between.

**Figure 3:**
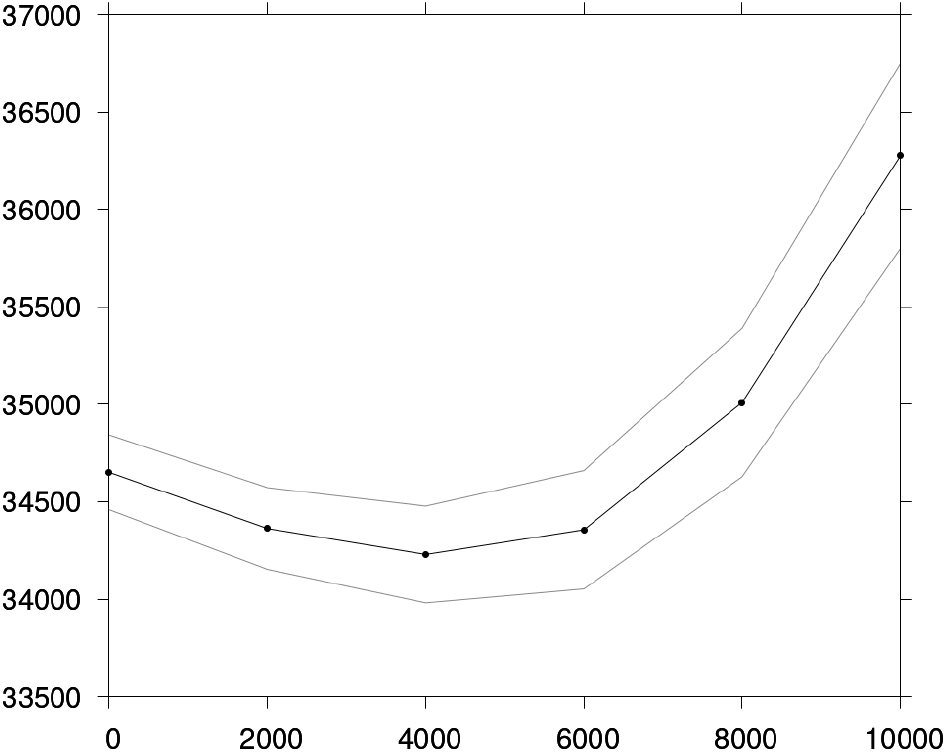
Ex ante expected reduction in symptomatic infected individual-days (y-axis) as a function of the number of administered single doses (x-axis) for *t*_min_ = 0 and *n*_max_ = 10,000. Thin lines: 95 % CI.

Importantly, the actual performance of policy *a*_0_ picked under uncertainty depends on the true values of uncertain parameters. In Figure 4, each dot corresponds to one of the 10,000 parameter draws shown in Figure 2. For each draw, we compare the performances of two policies: policy *a*_0_ consisting in administering a single dose to each of the 10,000 individuals in the population (x-axis), and the policy consisting in administering 5,000 double doses (y-axis). For most parameter values (dots below the *y* = *x* line), *a*_0_ does not perform better than administering 5,000 double doses. *a*_0_ is the best policy for only 48.51 % (95 % CI: 47.53 % – 49.49 %) of parameter values. Administering 5,000 double doses is the best policy for 50.96 % (95 % CI: 49.98 % – 51.94 %) of parameter values. Other policies perform better than the two shown here for 0.53 % (95 % CI: 0.41 % – 0.69 %) of parameter values.

**Figure 4:**
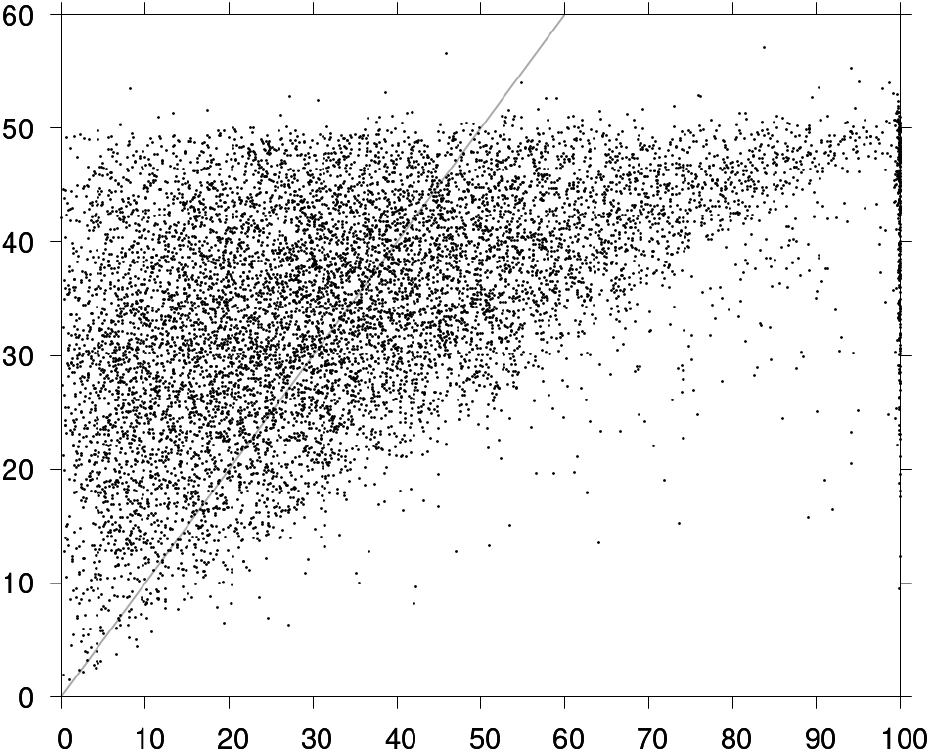
Reduction in symptomatic infected individual-days obtained with 10,000 single doses (x-axis, thousands) and with 5,000 double doses (y-axis, thousands) for 10,000 parameter draws. *t*_min_ = 0 and *n*_max_ = 10,000. Grey line: *y* = *x*.

Notice the difference between choosing *a*_0_ by maximizing the expected value over possible parameter values, and choosing the policy that is expected to be the best for most parameter values. With the latter approach, information about the relative performance of policies is lost. Still, Figure 4 shows that *a*_0_ will underperform for more than half of possible parameter values. The motivation of value of information analyses is to determine whether the additional benefit of picking the right policy knowing the true parameter values is worth the effort of investigating these true values.

We refer to Appendix D for additional comments on Figures 3 and 4.

### 3.2 Value of information under constraints

Let us assume that true parameter values can be known and the best policy picked for each particular parameter values. Then, still in the case *t*_min_ = 0 and *n*_max_ = 10,000, the expected burden reduction over possible parameter values obtained by picking the right policy knowing the true parameter values is 43,701.4 (95 % CI: 43,322.1 – 44,080.8) averted symptomatic infected individual-days. This is the expected ex post maximum value. EVPI is the difference between the expected ex post maximum value, and the ex ante maximum expected value: 43, 701.4 − 36, 274.5 = 7, 426.94 (95 % CI: 7,226.78 – 7,627.09) averted symptomatic infected individual-days. Recall that EVPI depends on prior parameter distributions, as shown in Appendix E. We give results for plausible prior distributions that correspond to a high EVPI in order to magnify the trade-offs we want to illustrate.

Table 2 shows EVPPI for subsets of uncertain parameters. Uncertainty is overwhelmingly driven by vaccine parameters, in particular the efficacy of single dose vaccination compared to no vaccination. Resolving uncertainty about *R*_0_, that is uncertainty about the spread of the disease without intervention, has comparatively little value. This may have implications in terms of how the decision problem is framed. In the present article, we focus on trade-offs between mutually exclusive options: resolving uncertainties, and vaccinating faster or with more doses. However, in some cases, vaccinating might allow to collect information about the vaccine’s properties. Whether this option should be retained depends largely on technical feasibility, and of course on political and ethical acceptability. This scenario is left outside the scope of the paper.

**Table 2:**
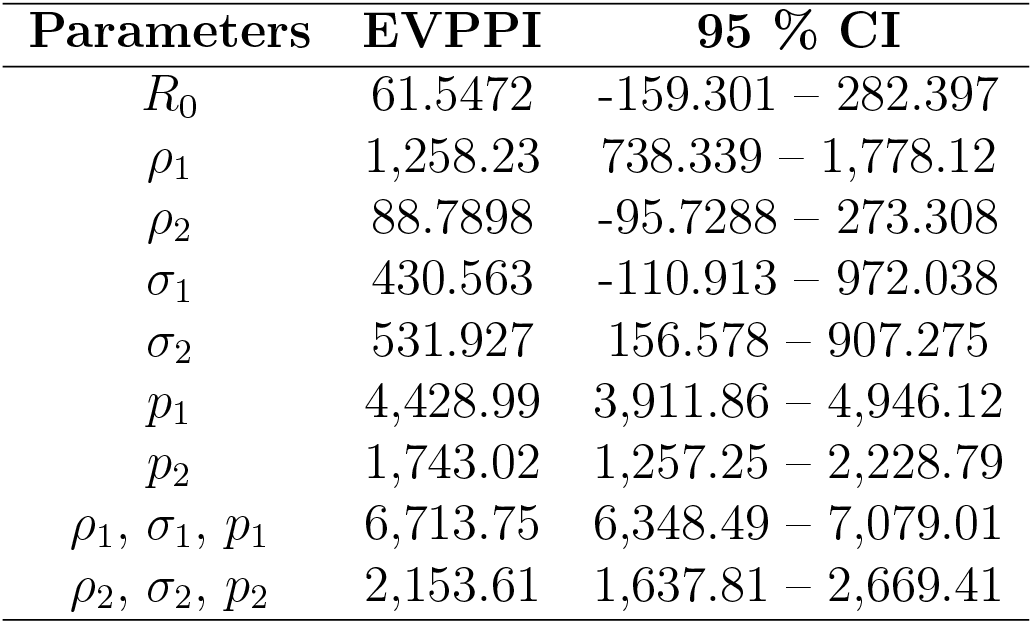
EVPPI in averted symptomatic infected individual-days for subsets of uncertain parameters in the case *t*_min_ = 0 and *n*_max_ = 10,000.

The results presented so far only concerned the case *t*_min_ = 0 and *n*_max_ = 10,000. Figure 5 shows the EVPI as a function of the set *A*(*t*_min_, *n*_max_) of feasible policies with variable constraints on the time of implementation *t*_min_ and number of available doses *n*_max_. Figures 5b and 5c are sectional views for *n*_max_ = 10,000 and *t*_min_ = 0 respectively of the heat map in Figure 5a. The EVPI is lower for later policy implementation dates *t*_min_ (Figure 5b, x-axis in Figure 5a). As time passes, the epidemic spreads and resolving uncertainties has less value. The number of available vaccine doses *n*_max_ is shown on the y-axis in Figure 5a and the x-axis in Figure 5c. When there is no vaccine dose available (*n*_max_ = 0), no individual can get vaccinated with or without information, and the EVPI is zero. With 20,000 available vaccine doses, each of the *N* = 10,000 individuals in the population can receive two doses, so that collecting information does not bring additional value and the EVPI is zero. The EVPI is maximal for *n*_max_ = 10,000 available vaccine doses. Here, 10,000 individuals can receive one dose or 5,000 individuals two doses. When less than 10,000 doses are available, the EVPI is lower because additional information cannot be fully taken advantage of due to a lack of doses. When more than 10,000 doses are available, the EVPI is lower because all individuals can receive a single dose and some of them a double dose, so the performance of policies picked based on additional information is closer to that of policies picked without resolving uncertainties.

**Figure 5:**
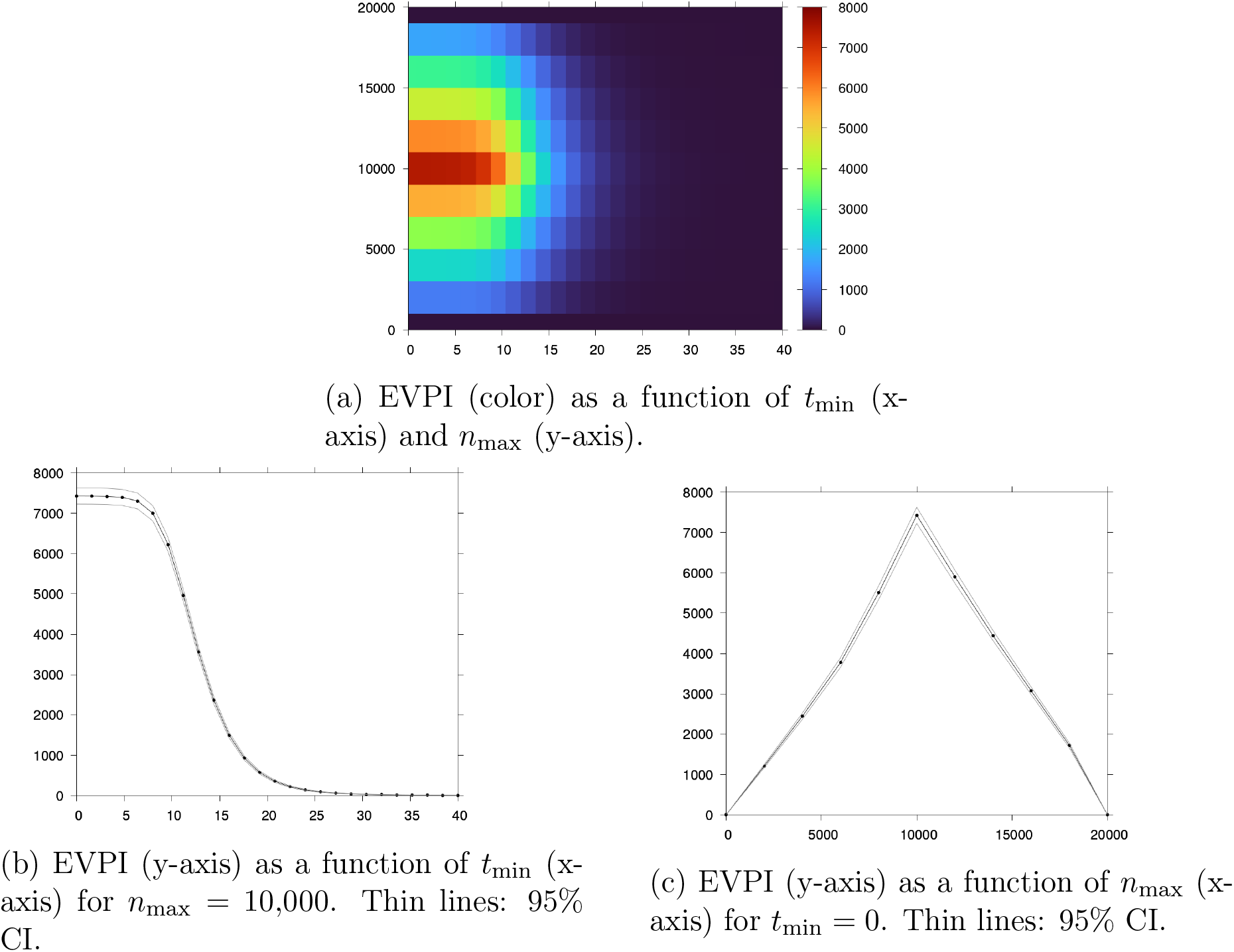
EVPI in averted symptomatic infected individual-days as a function of *A*(*t*_min_, *n*_max_).

### 3.3 Relaxing constraints v. resolving uncertainties

We illustrated how the set of available policy options, that is constraints on policies, has an influence on the expected value of resolving uncertainties. In order to show the trade-off between resolving uncertainties and relaxing policy constraints *t*_min_ and *n*_max_, we need to compute the ex ante maximum expected value and the expected ex post maximum value separately for different levels of constraint.

Figure 6 shows the ex ante maximum expected value and the expected ex post maximum value of control as a function of policy implementation date *t*_min_, for 10,000 available vaccine doses (see Appendix F for heat maps). For any given implementation time, more information is better than less information (EVPI is positive or null by definition). For a given level of information, vaccinating earlier is better than vaccinating later. This corresponds to the common intuition that early interventions prevent the subsequent spread of pathogens. However, the figure also shows that vaccinating at time 0 without additional information (dashed line) allows to avert the same number of symptomatic infected individual-days as vaccinating about 10 days later with perfect information (continuous line). Thus, starting from time 0 under uncertainty, collecting information is a better option than vaccinating at time 0 without additional information, as long as collecting information allows to resolve uncertainties in less than 10 days. The graph can also be read the other way round. Assume that vaccination was initially planned at time *t*_min_ = 10, e.g. because the 10,000 vaccine doses will only be ready at that time. In our scenario, it is better to spend additional resources to resolve uncertainties before *t*_min_ = 10 and vaccinate at *t*_min_ = 10 with perfect information, rather than spend them to accelerate vaccine production and bring vaccination forward (but without additional information), that is to relax the *t*_min_ constraint.

**Figure 6:**
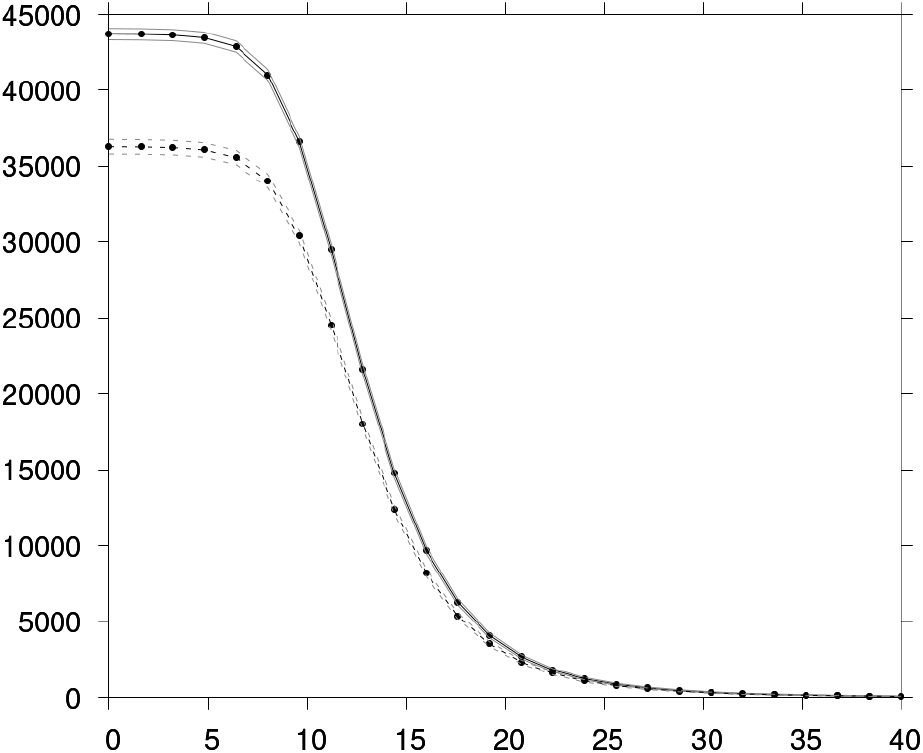
Ex ante maximum expected value (dashed line) and expected ex post maximum value (continuous line) as a function of *t*_min_ for *n*_max_ = 10,000. Thin lines: 95% CI.

Similarly, Figure 7 shows the ex ante maximum expected value and the expected ex post maximum value of control as a function of the number of available vaccine doses *n*_max_ for implementation date *t*_min_ = 0. For any number of available vaccine doses, more information is better than (or equivalent to, for *n*_max_ = 0 and *n*_max_ = 20,000) less information. For a given level of information, vaccinating with more doses is better than vaccinating with less doses. However, starting for instance from *n*_max_ = 10,000 available doses under uncertainty (dashed line), approximately the same number of averted symptomatic infectious individual-days can be achieved by resolving uncertainties (continuous line) or by acquiring approximately 2,000 more doses (dashed line), in other words by relaxing the *n*_max_ constraint. Thus, resolving uncertainties is a better option if it is less expensive than acquiring or producing 2,000 doses of vaccine. Finally, though this is a rather far-fetched scenario, up to 2,000 doses out of 12,000 could be sold if the benefit allowed to entirely resolve uncertainties.

**Figure 7:**
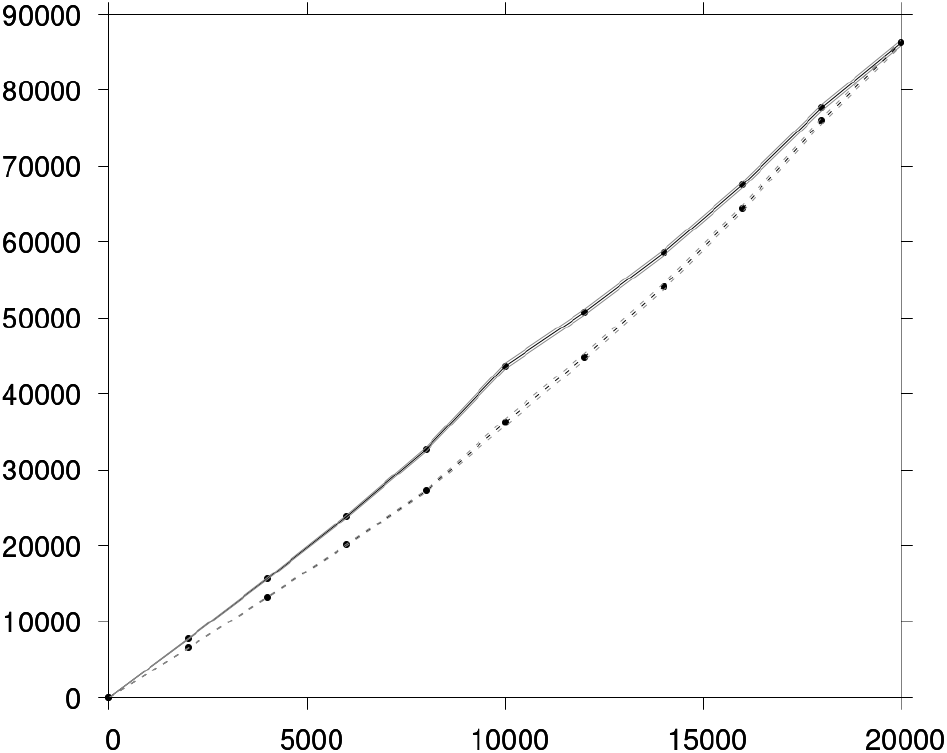
Ex ante maximum expected value (dashed line) and expected ex post maximum value (continuous line) as a function of *n*_max_ for *t*_min_ = 0. Thin lines: 95% CI.

## 4 Conclusion

EVPI is typically used to estimate the value of resolving uncertainties regarding an epidemic given a set of feasible control policies. In this article, we used an EVPI framework to explore the trade-off between resolving uncertainties and relaxing policy constraints, that is increasing the set of feasible control policies. We focused on two constraints: the implementation date, and the number of available vaccine doses. We asked respectively “is it better to use resources to bring vaccination forward or to resolve uncertainties?”, and “is it better to use resources to acquire more vaccine doses or to resolve uncertainties?”. We first reminded the rationale for value of information analyses by illustrating how decisions made under uncertainty can underperform, then we showed how EVPI depends on policy constraints. Finally, we compared the expected ex post value of control (after uncertainties are resolved) with the ex ante expected value of control (before uncertainties are resolved) for different levels of policy constraints in order to show how relaxing constraints compares to resolving uncertainties.

Similar trade-offs have been presented in previously published studies, but most often implicitly in the context of multi-stage epidemic control. Other studies discussed such trade-offs explicitly, but only for a given type of policy constraint, not over the full range of constraints, or in very special cases. We believe that the trade-offs between relaxing policy constraints and resolving uncertainties deserved a more thorough discussion at a more abstract level.

The main results of the paper are based on EVPI. We must emphasize that EVPI is only one value of information metric among others. It assumes that uncertainties can be entirely resolved, which is unlikely to be the case in real life. Thus it only provides upper bounds on the value of collecting information. Besides, as noted by other authors [1], the time and effort required to collect information may depend on the parameters. The success of data collection may also be uncertain. In practice, we expect value of *sample* information metrics such as the expected value of sample information (EVSI) to be of use, since information is usually collected only partially e.g. through sampling (see for example Cipriano and Weber [16]). We left the value of sample information outside the scope of this article to avoid going into the technical specifics of information collection, e.g. through lab experiments, clinical studies, or from real-life data depending on the case. Notice also that for a given sample size, an EVSI analysis would give results qualitatively similar to our EVPI analysis. To remain general, we did not look into the details of the cost of data collection and of vaccination campaigns, including the fact that producing vaccine doses may take time. These costs can be critical in practice and will depend on each specific case. Overlooking the cost details here does not make our results any less relevant since we give results in terms of upper bounds of the value of information. We also assumed that collection of information and, say, vaccination with more doses, are two mutually exclusive uses of resources. Yet vaccinating more could actually bring information about the vaccine’s properties [18], and our EVPPI analysis suggests that such scenarios might indeed be worth consideration. Independently of EVPPI considerations, it is likely that uncertainty about some parameters can only be resolved “by doing”, that is by vaccinating. This would be the case, for instance, of the number of individuals accepting of refusing vaccination (this parameter was not considered in the present article). Like EVSI, we left these scenarios aside because they mostly concern uncertainty on some of the parameters (the vaccine’s properties), and because they raise the question of *how* to collect information in practice and to infer information from specific data, which is not the point of our article (we merely look at the value of information).

Importantly, EVPI depends on modeling choices, some of which involve not only scientific but also political and ethical considerations. Determining the possible values of uncertain parameters and their likelihood is primarily a scientific or technical issue. The choice of a policy performance metric, by contrast, is also a political and ethical question: consider for instance the difference between maximizing the number of avoided deaths and saving a maximum of life-years. The same applies to the chosen objective. In this, we assumed that the objective is to maximize the expected value over uncertain parameter values. The value of information framework can be adapted to alternative objectives such as minimizing the probability of extreme adverse events (see the examples provided by Shea et al. [17]).

We used a generic transmission model and a generic value of information metric to make a very general argument. In this article, the SIR model, EVPI, and fast vaccination of the population on a given date, should be understood as minimal working examples of a disease transmission model, value of information metric, and vaccination intervention respectively. The approach presented here is entirely based on numerical simulations of markovian models and is therefore highly versatile. Implementing the appropriate compartmental model, using the right value of information metric, or modeling a vaccination campaign with, say, a ramp-up phase and target age-groups is a mere problem of engineering and computing. The trade-offs illustrated in this article will still hold, although they will most likely be trivial in some special cases, e.g. if EVPI is zero or close to zero for any level of policy constraints.

Hitting fast and hard is commonly regarded as a sensible policy in the face of an epidemic. Using resources to acquire information instead, it might be argued, raises issues of ethical and political acceptability somewhat reminding of the colloquial trolley problem. Yet arguably any political or ethical constraint (or objective) can be included in a value of information analysis e.g. by choosing a performance metric and setting an objective (maximizing the expected value over uncertain parameters, minimizing the probability of adverse outcome, etc.). Thus, waving information collection aside *a priori* and without further consideration can hardly be passed off as a deliberate ethical or political stance. It rather indicates flawed decision making.

## Data Availability

All data produced in the present study are available upon reasonable request to the authors

## Appendix

### A Compartmental model

Let *S*_*i*_ and *I*_*i*_ denote the number of susceptible and infectious individuals who received *i* ∈ {0, 1, 2} doses of vaccine, and *R* the number of recovered individuals. We assume a closed population of *N* individuals.

Individuals get infected with contact rate *β* and they recover at rate *γ*. The basic reproduction number is *R*_0_ = *β/γ*. Individuals who received *i* ∈ {1, 2} vaccine doses are less susceptible by a factor *σ*_*i*_ and less infectious by a factor *ρ*_*i*_ compared to individuals who received *i* − 1 doses. We assume that vaccination is instantaneous and that there is no waning of vaccine protection. The epidemic dynamics is described by Equations (1)–(7).

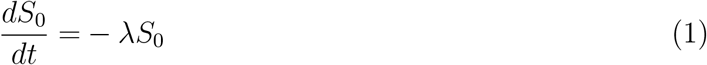

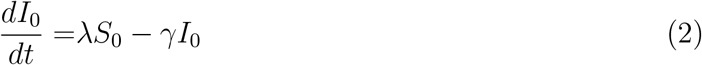

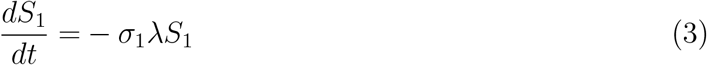

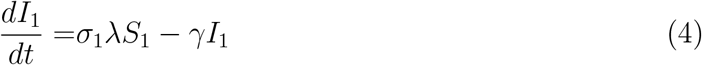

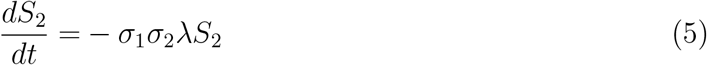

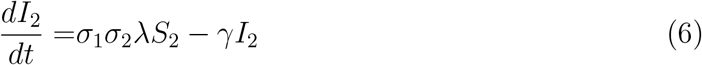

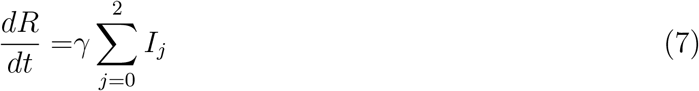

where

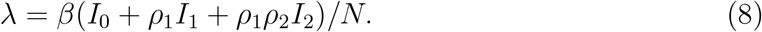

Let *p*_*i*_ the relative probability of symptomatic infection for infected individuals who received *i* ∈ {1, 2} doses of vaccine, compared to individuals who received *i* − 1 doses. We assume that all infected individuals who did not vaccinate become symptomatic. The incidence rates of symptomatic cases among unvaccinated individuals is 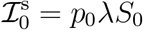. Among individuals who received 1 and 2 doses of vaccine, incidence rates of symptomatic cases are 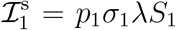 and 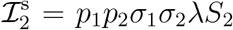, and the incidence rates of asymptomatic cases are 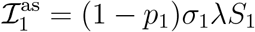 and 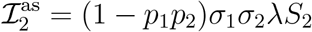.

**Table App-1:**
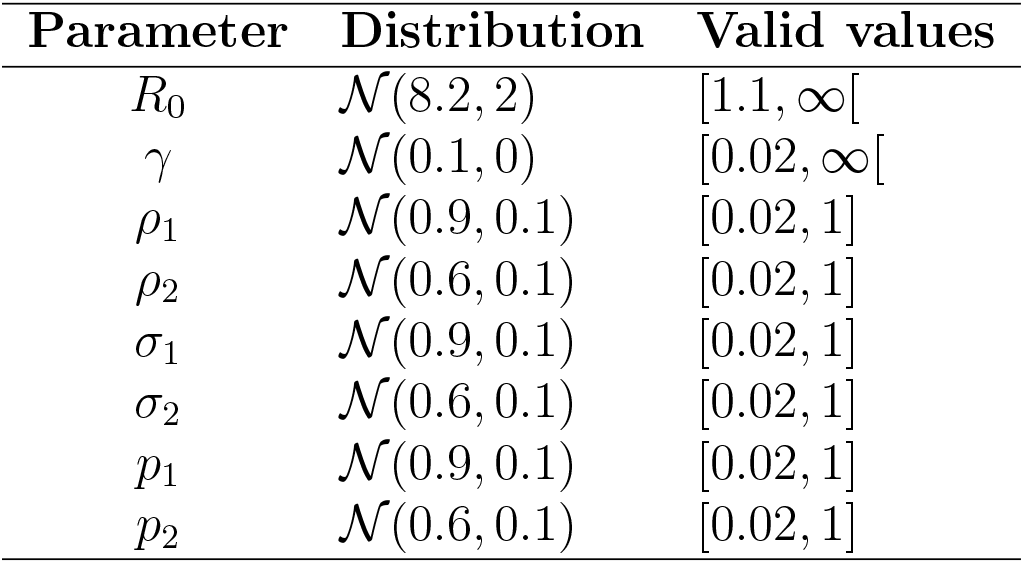
Prior distribution for parameters. 𝒩 (*μ, σ*^2^): normal distribution with mean *μ* and variance *σ*^2^. Distributions are truncated with support shown in the last column.

| Parameter | Distribution | Valid values |
| --- | --- | --- |
| $R_0$ | $\mathcal{N}(8.2, 2)$ | $[1.1, \infty[$ |
| $\gamma$ | $\mathcal{N}(0.1, 0)$ | $[0.02, \infty[$ |
| $\rho_1$ | $\mathcal{N}(0.9, 0.1)$ | $[0.02, 1]$ |
| $\rho_2$ | $\mathcal{N}(0.6, 0.1)$ | $[0.02, 1]$ |
| $\sigma_1$ | $\mathcal{N}(0.9, 0.1)$ | $[0.02, 1]$ |
| $\sigma_2$ | $\mathcal{N}(0.6, 0.1)$ | $[0.02, 1]$ |
| $p_1$ | $\mathcal{N}(0.9, 0.1)$ | $[0.02, 1]$ |
| $p_2$ | $\mathcal{N}(0.6, 0.1)$ | $[0.02, 1]$ |

### B Prior distributions for parameters

#### C Vaccination policies

##### C.1 Definition

We define a vaccination session as a tuple (*t, d, d*′, *n*) where *t* is the vaccination date of the target population, *d* is the number of doses received by individuals in the target population prior to the vaccination session, *d*′ *> d* is the number of doses received by each individual in the target population after the vaccination session, and *n* is the total number of doses administered in the target population. For instance, (2, 0, 2, 4000) reads: at time *t* = 2, give *d*′ − *d* = 2 doses to *n/*(*d*′ − *d*) = 2, 000 individuals that have not received any dose previously (*d* = 0) – a total of *n* = 4000 doses of vaccine are administered for this session.

Notice that a target population is only defined by the number *d* of previously received doses. For simplicity, individuals to be vaccinated during the session are randomly drawn in the target population. In practice, though, vaccination campaigns could be made more efficient by using information on who is symptomatically infected and who recovered from a symptomatic infection and targeting individuals accordingly.

In general, a vaccination policy is made up of several consecutive or simultaneous vaccination sessions. Vaccination session and policies are subject to several feasibility constraints (e.g. there should be enough individuals in target populations). These constraints are given formally in Appendix C.2. In this article, we consider the sets of feasible alternative policies

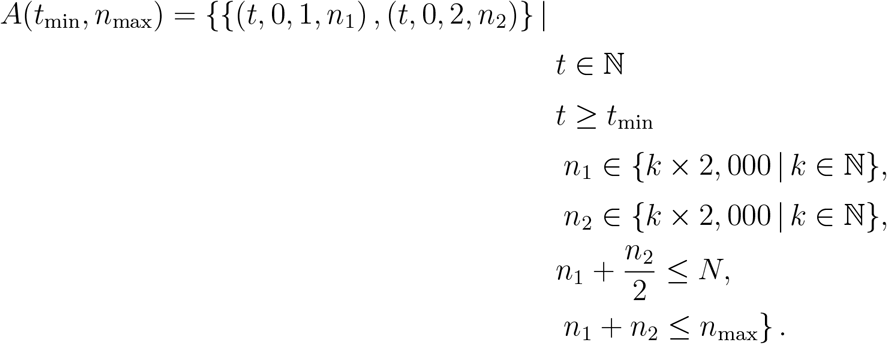

where *t*_min_ is the earliest date at which the vaccination sessions can be implemented and *n*_max_ is the maximum number of vaccination doses administered. Trivially, vaccination is implemented at time *t*_min_ in our case.

##### C.2 Feasibility

Let 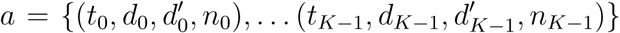 a vaccination policy made up of *K* vaccination sessions. For the sake simplicity, we restrict to the case where time is discontinuous – *i*.*e*. 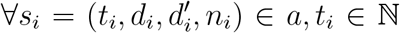 and where individuals can received up to 2 vaccine doses – *i*.*e*. 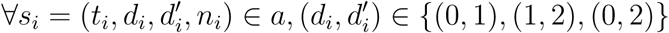.

For the sake of simplicity and with a slight abuse of notation, we say that (*t, d, d*′) ∉ *a* if ∀*n* ∈ ℕ, (*t, d, d*′, *n*) ∉*a*. We define 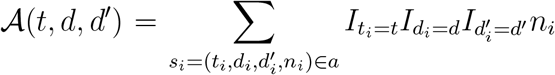 where *I* is the indicator function. Hence, 𝒜 (*t, d, d*′) is the number of doses administered at time *t* to individuals who previously received *d* doses and currently receive *d*′ − *d* doses.

We call 𝒩 (*d, t*) with *d* ∈ {0, 1, 2} and *t* ∈ {−1} ∪ ℕ, the number of individuals who received *d* doses before date *t*. Formally, we define 𝒩 recursively with:

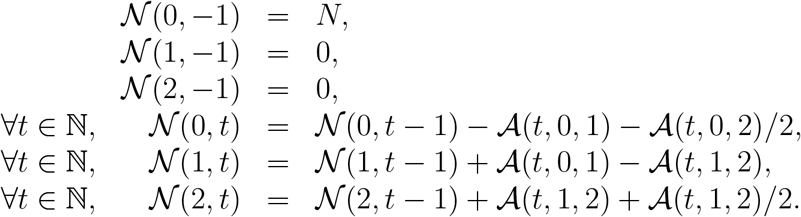

We say that *a* is feasible if the following constraint is met: ∀*t* ∈ ℕ, 𝒩 (0, *t*) ≥ 0, 𝒩 (1, *t*) ≥ 0, 𝒩 (2, *t*) ≥ 0. Notice that by definition ∀*t* ∈ ℕ, 𝒩 (0, *t*) + 𝒩 (1, *t*) + 𝒩 (2, *t*) = *N* so that it is unnecessary to check that each vaccination state subpopulation is inferior to *N*.

### D Additional comments: decision under uncertainty

The curve in Figure 3 is convex:

- For high numbers of protected individuals (e.g. 8,000 single doses and 1,000 double doses), the burden reduction is mostly driven by the number of protected individuals, so it is marginally better to increase that number with more single doses and less double doses.
- For low numbers of protected individuals (e.g. 2,000 single doses and 4,000 double doses), the burden reduction is mostly driven by the higher efficacy of double doses, so it is marginally better to increase the number of double doses even by decreasing the total number of protected individuals.

The distribution of dots in Figure 4 can be explained as follow. Recall that symptomatically infected individuals are symptomatic during 10 days on average (*γ* = 0.1) and the population is *N* = 10,000 individuals, so at most 100,000 symptomatic infected individual-days can be averted. The direct protection offered by vaccination (reduction of symptom probability) dominates indirect protection (reduction of transmission). For this reason, administering 5,000 double doses (y-axis) allows to avert at most 50,000 symptomatic infected individual-days for most parameter values. The few dots lying above the line *y* = 50, 000 correspond to parameter values such that *R*_0_ is low enough for indirect protection to have an impact. Most points are above the line *y* = 0.5*x* for the same reason, that the direct vaccine protection dominates indirect effects in our scenario. Only 5,000 individuals are treated with double doses (y-axis) versus twice as many with single doses (x-axis). Dots that are close to the line *y* = 0.5*x* correspond to parameter values such that double dose vaccination does not significantly reduce symptom probability compared to single dose vaccination. In this case, the burden reduction with single doses is about twice the burden reduction with double doses simply because twice as many individuals are protected. Most points are above the line because double dose vaccination offers additional direct protection for most parameter values. If indirect protection played a significant role, we would not see a frontier at *y* = 0.5*x*. Points below the line *y* = 0.5*x* correspond to parameter values such that indirect effects are significant with one dose of vaccine. In such cases, vaccinating half as many individuals with a marginally more efficient double vaccine dose offers less that half the benefit of single dose vaccination of twice as many individuals.

**Table App-2:**
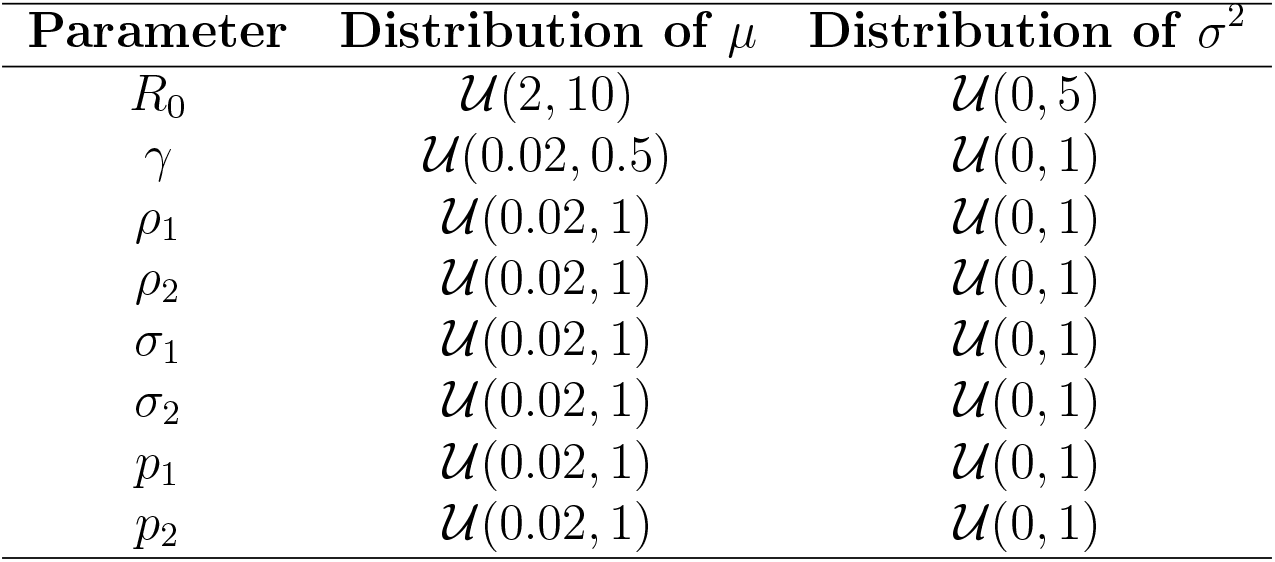
Distributions of prior distributions’ parameters *μ* and *σ*^2^. 𝒰 (*a, b*): uniform distribution on [*a, b*]

| Parameter | Distribution of $\mu$ | Distribution of $\sigma^2$ |
| --- | --- | --- |
| $R_0$ | $\mathcal{U}(2, 10)$ | $\mathcal{U}(0, 5)$ |
| $\gamma$ | $\mathcal{U}(0.02, 0.5)$ | $\mathcal{U}(0, 1)$ |
| $\rho_1$ | $\mathcal{U}(0.02, 1)$ | $\mathcal{U}(0, 1)$ |
| $\rho_2$ | $\mathcal{U}(0.02, 1)$ | $\mathcal{U}(0, 1)$ |
| $\sigma_1$ | $\mathcal{U}(0.02, 1)$ | $\mathcal{U}(0, 1)$ |
| $\sigma_2$ | $\mathcal{U}(0.02, 1)$ | $\mathcal{U}(0, 1)$ |
| $p_1$ | $\mathcal{U}(0.02, 1)$ | $\mathcal{U}(0, 1)$ |
| $p_2$ | $\mathcal{U}(0.02, 1)$ | $\mathcal{U}(0, 1)$ |

### E Sensitivity of EVPI to prior distributions

We compute EVPI for 2000 prior distributions of uncertain parameters (see Table App-1 for the baseline prior distributions). The 2000 prior distributions are generated by drawing parameters *μ* and *σ*^2^ in the uniform distributions shown in Table App-2. Figure App-1 shows the 2000 EVPIs sorted by in increasing order.

**Figure App-1:**
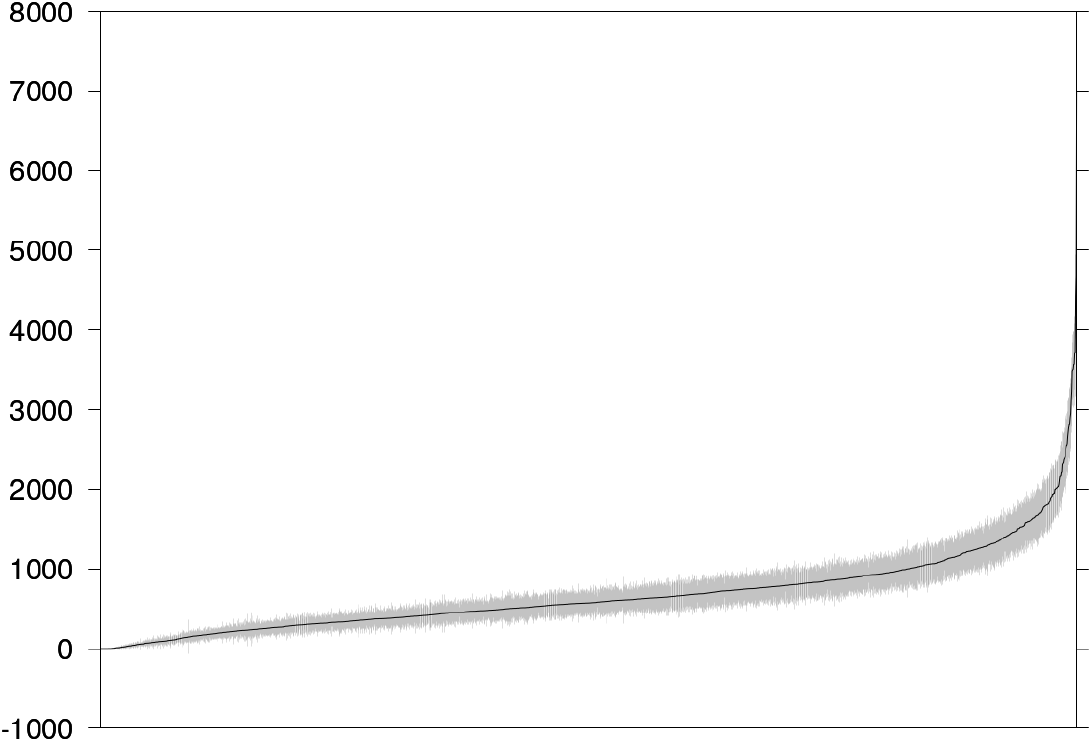
EVPIs for 2000 randomly generated prior distributions of random parameters. Grey: 95% CI.

### F Additional figures: ex ante maximum expected value v. expected ex post maximum value

Figure App-2 shows the ex ante maximum expected value max 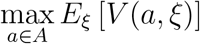 and the expected ex post maximum value 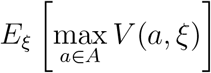 as a function of *A*(*t*_min_, *n*_max_).

**Figure App-2:**
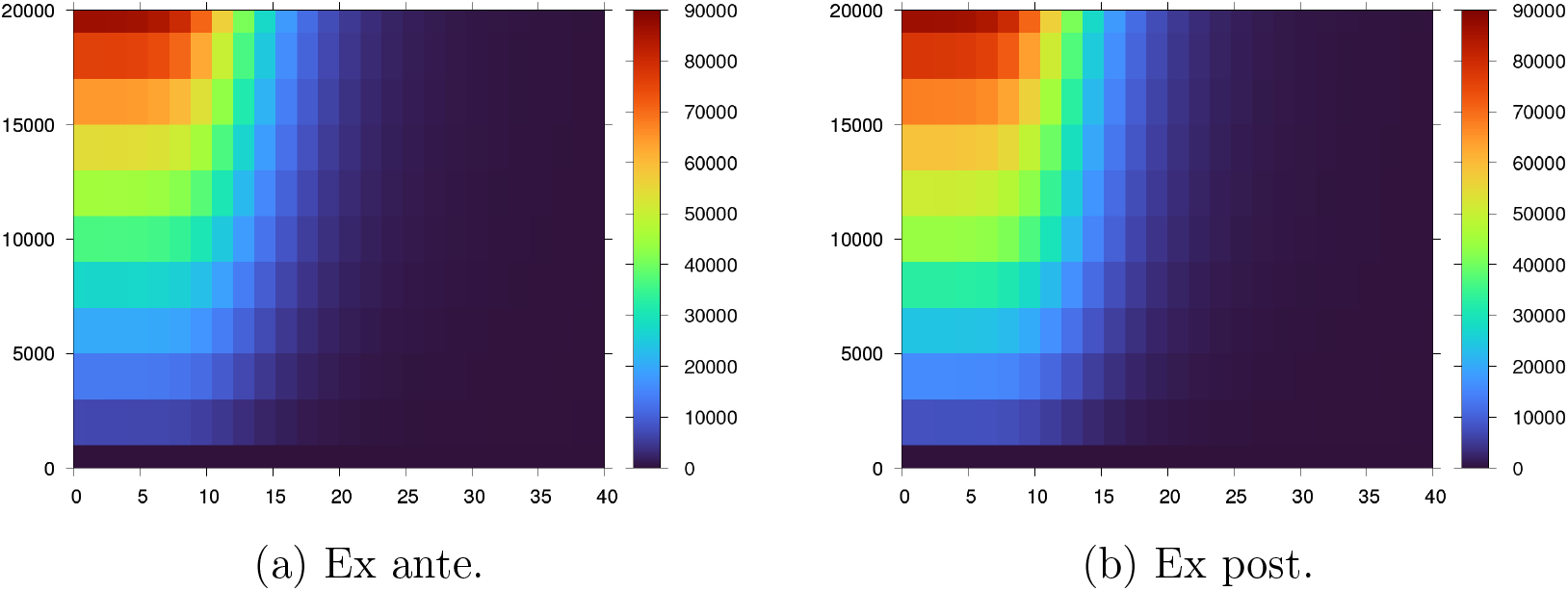
Ex ante maximum expected value and expected ex post maximum value as a function of *A*(*t*_min_, *n*_max_). x-label: policy implementation date *t*_min_. y-label: *n*_max_.

## Footnotes

1 So the same information is used to choose a policy and to test its robustness. Notice that the chosen policy already takes that information into account, hence a tautology. Contrast this with value of information approaches: a policy chosen under uncertainty is deemed robust if knowing the true value of parameters does not allow to pick a much better policy.

